# MSPTDfast: An Efficient Photoplethysmography Beat Detection Algorithm

**DOI:** 10.1101/2024.07.18.24310627

**Authors:** Peter H Charlton, Jonathan Mant, Panicos A Kyriacou

## Abstract

Beat detection is a key step in the analysis of photo-plethysmogram (PPG) signals. The ‘MSPTD’ algorithm was recently identified as one of the most accurate beat detection algorithms, but its current open-source implementation is substantially more computationally expensive than other leading algorithms such as ‘qppgfast’. The aim of this work was to develop a more efficient, open-source implementation of the ‘MSPTD’ algorithm. Five potential improvements were identified to increase efficiency. Each potential improvement was evaluated in turn, and an optimal algorithm configuration named ‘MSPTDfast’ was developed which incorporated all of the improvements found to reduce algorithm execution time whilst not substantially reducing the accuracy of beat detection. Performance was assessed using data collected from young adults during a lunchbreak in the PPG-DaLiA dataset. The data consisted of wrist PPG signals acquired using an Empatica E4 device, alongside simultaneous ECG signals from which reference heartbeat timings were obtained. ‘MSPTDfast’ was found to be substantially more efficient than ‘MSPTD’ (a reduction in execution time of 72.3%), with minimal difference in beat detection accuracy (F_1_-score 87.8% vs. 87.7%). In addition, the performance of ‘MSPTDfast’ was much closer to that of the state-of-the-art ‘qppgfast’ algorithm than the ‘MSPTD’ algorithm, with a comparable F_1_-score (87.4% vs. 87.7%), and an execution time which was only 19.2% longer than that of ‘qppgfast’ (vs. 330.8% longer for ‘MSPTD’). In conclusion, ‘MSPTDfast’ is an efficient and accurate open-source PPG beat detection algorithm with a substantially faster execution time than ‘MSPTD’. It is available under the permissive MIT licence.

## 1. Introduction

Photoplethysmography, an optical sensing technology, is now widely used in physiological measurement. Photoplethysmography sensors are incorporated into many wearable devices such as smartwatches and smart rings, and photoplethysmogram (PPG) signals can also be acquired by everyday devices such as smartphones and webcams. A plethora of physiological parameters can be estimated from PPG signals, such as heart rate, heart rhythm, respiratory rate, and blood pressure [1].

A key step in PPG signal processing is beat detection: identifying individual pulse waves corresponding to heart beats. Several PPG beat detection algorithms are openly available, of which *‘MSPTD’* and *‘qppg’* have recently been found to be most accurate [2]. Since these algorithms are strong candidates for PPG analysis, it is important to develop efficient open-source implementations of them. Previously, an efficient version of *‘qppg’* has been developed named *‘qppgfast’* [3]. In addition, there have been efforts to develop efficient implementations of *‘MSPTD’*, with *‘MSPTD’* itself being a more efficient version of the original *‘AMPD’* algorithm [4, 5], and further refinements having been proposed [6]. However, the open-source implementation of the *‘MSPTD’* algorithm [7] is substantially more computationally expensive than the *‘qppgfast’* algorithm, limiting its potential utility.

The aim of this work was to develop a more efficient, open-source implementation of the *‘MSPTD’* algorithm for photoplethysmography beat detection. A series of potential algorithm improvements were systematically evaluated, and a final configuration was tested on wrist PPG signals from a wearable device.

## 2. Methods

### 2.1. The MSPTD Algorithm

The MSPTD algorithm has been described previously, so is only briefly described here (see [4] for the original description, [5] for a description of the original algorithm on which it was based, and [6] for a further explanation).

The algorithm consists of the following steps, as implemented in [7]:

- Segment the signal into 6s windows, with 20% overlap.
- Detrend the signal.
- Produce *‘local maxima scalograms’* (LMSs) for peaks and onsets, where an LMS is a matrix of logical values where each column corresponds to a sample in the signal, and each row indicates whether or not that sample is higher (for peaks) or lower (for onsets) than its neighbours at a particular scale (*i*.*e*. a particular separation between the sample and its neighbours on either side). Consider all scales from 1 sample separation to a separation of *N*/2, where *N* is the number of samples in the signal.
- Identify the scale, *γ*, with the most local maxima (*i*.*e*. peaks) or minima (*i*.*e*. onsets).
- Truncate the LMS(s) to remove rows corresponding to scales larger than *γ*.
- Identify peaks (or onsets) as columns in the LMSs where all values are true, indicating that those signal samples were local maxima (or minima) at all considered scales.
- Refine the location of each identified peak (or trough) by searching for the highest (or lowest) sample within 50 ms either side of the original location.

### 2.2. Potential Algorithm Improvements

We identified the following potential improvements to the current *‘MSPTD’* implementation:

- Calculate only one LMS, corresponding to either pulse wave peaks or onsets, instead of the original two LMSs.
- Vectorise the LMS calculation method to avoid its computationally expensive nested for loops, as proposed in [6].
- Reduce the LMS size by excluding scales corresponding to frequencies below a minimum heart rate, *HR*_*min*_.
- Downsample the PPG signal prior to beat detection, thus reducing the size of the LMS.
- Adjust the PPG window duration to be shorter or longer than the original value of 6 s.

### 2.3. Evaluating Potential Improvements

For each potential improvement we identified different options, as summarised in Table 1. Some options deserve comment: (i) whilst calculating two LMSs to identify both pulse wave peaks and onsets is expected to be more computationally expensive than only identifying either peaks or onsets, it may improve the accuracy of beat detection; (ii) minimum sampling frequencies of 10, 20 and 30 Hz were chosen as most content in PPG signals is thought to be below 8-25 Hz [8]; (iii) shorter PPG window durations will reduce the size of the LMS, but may also decrease the accuracy of beat detection.

**Table 1.** Evaluated algorithm configurations, where * indicates a default option.

| Potential improvement | Options |
| --- | --- |
| LMSs to calculate | peaks; onsets;<br>peaks and onsets * |
| LMS calculation method | nested for loops *;<br>vectorised approach |
| LMS scales used | $N/2$ *;<br>only scales $>HR_{min}$, with<br>$HR_{min} \in \{30, 40\}$ bpm. |
| Sampling frequency (Hz) | 10; 20; 30; original * |
| Window duration (s) | 4; 6; 8 *; 10 |

The performance of a refined *‘MSPTD’* algorithm was assessed when using each possible option whilst all others were held at default values. This assessment was performed using the *‘ppg-beats’* framework for assessing PPG beat detection algorithms [2], which produces two metrics: (i) the *F*_1_-score, indicating the accuracy of beat detection, where beats are deemed accurate if they are within ±150 ms of reference ECG beats; and (ii) the algorithm execution time, the time taken to run the algorithm on a computer (in this case a MacBook Air M1 2020 without parallelisation), expressed as a percentage of the PPG signal duration.

The publicly available PPG-DaLiA dataset was used [9]. It contains wrist PPG signals acquired using the Empatica E4 device, alongside chest ECG signals, collected from 15 subjects in a protocol of daily living activities. The subjects were aged a median (lower-upper quartiles) of 28 (24–36) years, included three females, and had skin types on the Fitzpatrick scale of: 2 (1 subject),3 (11 subjects), and 4 (3 subjects) [2]. We used the subset of data collected during a lunch break, as it has previously been found to be suitably challenging for PPG beat detection [2]. This subset has a duration of 32.4 (28.7-37.2) minutes.

### 2.4. Designing and Evaluating *‘MSPTDfast’*

The *‘MSPTDfast’* algorithm was designed by selecting each configuration option which provided the shortest execution time whilst maintaining a reasonably high *F*_1_-score (a subjective process). Its performance was compared to two state-of-the-art algorithms: *‘MSPTD’* [7] and *‘qppgfast’* [3]. This evaluation was performed in MATLAB (The Mathworks, Natick, MA, USA).

## 3. Results

### 3.1. Potential Improvements

Figure 1 panels (a)-(e) show how the execution time (in blue) and and *F*_1_-score (in red) varied when using each option for each potential improvement.

**Figure 1.**
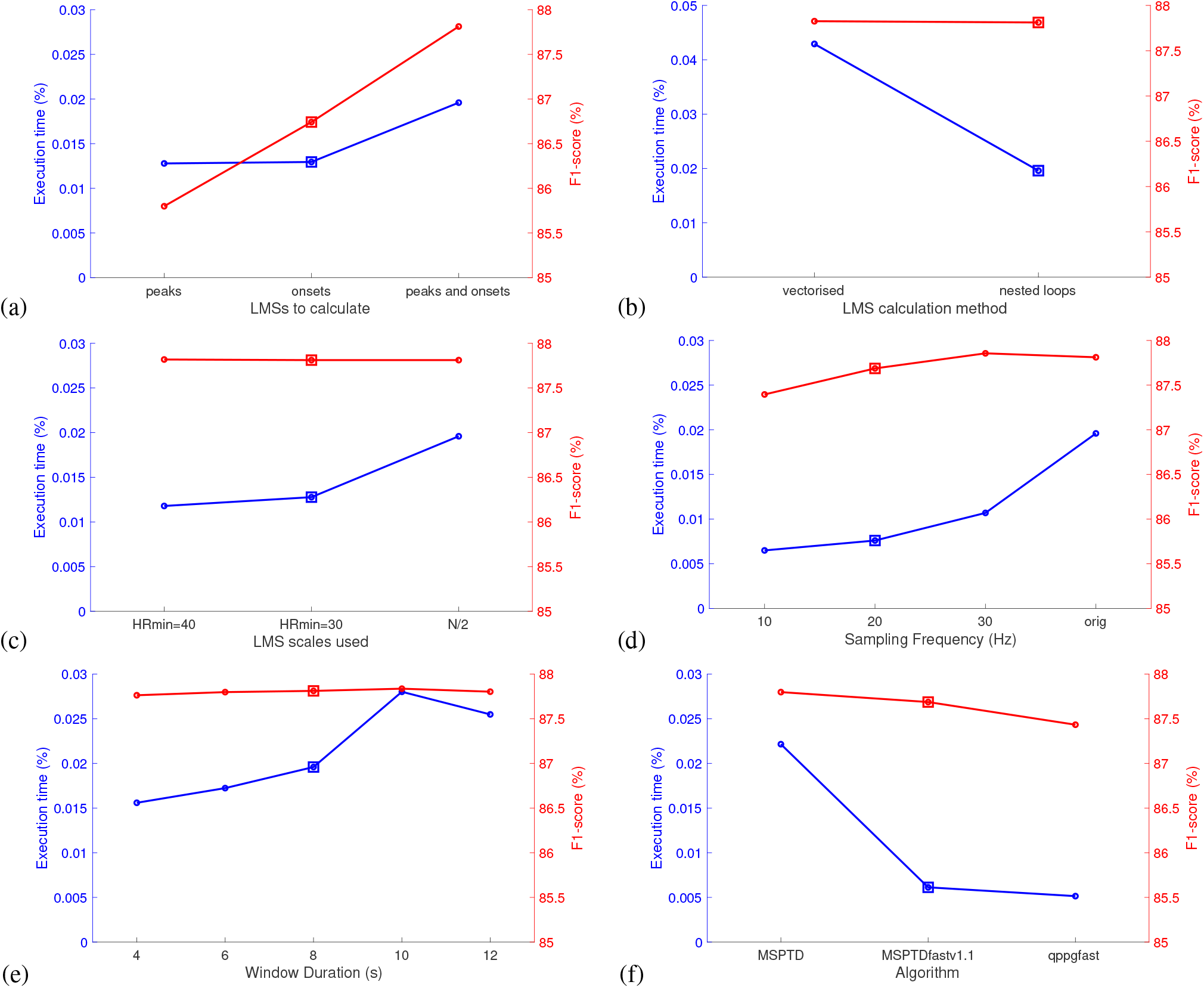
The performance of different algorithm configurations: (a)-(e) show performance when using different potential improvements, where squares indicate the configurations used in *‘MSPTDfast’* ; and (f) shows the performance of *‘MSPTDfast’* alongside two state-of-the-art algorithms - *‘MSPTD’* and *‘qppgfast’*.

Calculating only a single LMS corresponding to either peaks or onsets resulted in substantial reductions in execution time of 34.7% or 33.9% respectively, compared to calculating LMSs for both peaks and onsets (see (a)). This was accompanied by reductions in *F*_1_-score of 2.0% or 1.1% respectively. The original ‘peaks and onsets’ option was selected for *‘MSPTDfast’* as the moderate reduction in execution time provided by the other methods was not considered worthwhile when accompanied by a reduction in *F*_1_-score.

Using a vectorised approach to calculating the LMSs substantially increased execution time in our implementation (see (b)), and therefore the original ‘nested loops’ approach used in *‘MSPTD’* was retained for *‘MSPTDfast’*.

Reducing the number of LMS scales substantially reduced the execution time by 39.8% (with *HR*_*min*_ = 30) or 34.8% (with *HR*_*min*_ = 40) (see (c)). Whilst no reduction in *F*_1_-score was observed when using either *HR*_*min*_ value, we selected the more conservative *HR*_*min*_ = 30 bpm for *‘MSPTDfast’* to retain accuracy at low heart rates.

Reducing the sampling frequency substantially reduced execution time, with values of 30, 20, and 10 Hz reducing execution time by by 45.4%, 61.2% and 66.8% respectively (see (d)). These were accompanied by reductions in *F*_1_-score of 0.0%, 0.1% and 0.4%. Therefore, *‘MSPTDfast’* was configured to downsample signals to 20 Hz.

Reducing the window duration generally reduced execution time, accompanied by a slight reduction in *F*_1_-score (see (e)). A duration of 8 seconds was selected for *‘MSPTDfast’* in preference to even shorter durations which it was thought could reduce accuracy at lower heart rates.

### 3.2. The MSPTDfast Algorithm

Figure 1 (f) shows the performance of the *‘MSPTDfast’* algorithm alongside two state-of-the-art algorithms: *‘MSPTD’* and *‘qppgfast’*. The new *‘MSPTDfast’* algorithm had a execution time of approximately three-tenths (27.7%) of the *‘MSPTD’* algorithm, indicating a greater than three-fold reduction in execution time. This was achieved with only a very small reduction in *F*_1_-score of 0.1% (87.8% vs. 87.7%). In comparison to *‘qppgfast’*, *‘MSPTDfast’* had a longer execution time (19.2% longer) and a comparable *F*_1_-score (87.4% vs. 87.7%).

## 4. Discussion

In this study we developed *‘MSPTDfast’*, an efficient and accurate open-source algorithm for PPG beat detection. *‘MSPTDfast’* incorporates the following advances to improve efficiency compared to the original *‘MSPTD’* algorithm: the size of the 2D LMS matrix is substantially reduced in both directions by downsampling the PPG signal and reducing the number of scales over which beats are detected. In addition, an optimal window duration was used. The algorithm’s execution time was reduced by 72.3% in comparison to *‘MSPTD’* whilst retaining beat detection accuracy. The performance of the new *‘MSPTDfast’* algorithm is now much closer to that of the state-of-the-art *‘qppgfast’* algorithm (achieving similar accuracy, albeit with a 19.2% longer execution time) than the *‘MSPTD’* algorithm (with a 330.8% longer execution time).

The improved efficiency was mostly achieved by reducing the time spent on LMS calculation, which is the most computationally expensive part of the *‘MSPTD’* algorithm. However, the previously proposed improvement achieved by vectorising the LMS calculation rather than using nested for loops [6] was not successfully reproduced in this study. This may represent a shortcoming in our implementation of this approach. To our knowledge the previously proposed implementation is not openly available, making it difficult to reproduce this exactly.

A key limitation to this study is that *‘MSPTDfast’* was only assessed on a single, small dataset. Therefore, it is not yet clear whether it will generalise well to other datasets. For this reason we denote the current version of *‘MSPTDfast’* as ‘v.1.1’, in the hope that either ourselves or others will improve it further in the future.

The new *‘MSPTDfast’* algorithm complements *‘qppgfast’*. First, it is available under the permissive MIT license, whereas *‘qppgfast’* is available under the copyleft GNU General Public License. Second, *‘MSPTDfast’* uses a general approach which has been applied to disparate problems from astrophysics to chaos theory [5], with minimal tailoring for the PPG. In contrast, the *‘qppg’* approach was designed for cardiovascular pulse wave signals [10].

## 5. Conclusion

*‘MSPTDfast’* is an efficient and accurate open-source PPG beat detection algorithm with a substantially faster execution time than the *‘MSPTD’* algorithm on which it is based, and its execution time is much closer to that of the state-of-the-art *‘qppgfast’* algorithm. It is available at [7].

## Data Availability

This study used openly available data that were originally located at: https://doi.org/10.24432/C53890

https://doi.org/10.5281/zenodo.13121283

## Acknowledgments

This study is funded by the British Heart Foundation [FS/20/20/34626]. For the purpose of open access, the author(s) has applied a Creative Commons Attribution (CC BY) license to any Accepted Manuscript version arising.

